# Premenstrual disorders and risk of sick leave and unemployment: a prospective cohort study of 15,857 women in Sweden

**DOI:** 10.1101/2024.12.03.24318399

**Authors:** Hang Yu, Yihui Yang, Elgeta Hysaj, Alicia Nevriana, Sara Hägg, Unnur Anna Valdimarsdóttir, Elizabeth Bertone-Johnson, Donghao Lu

**Affiliations:** Unit of Integrative Epidemiology, Institute of Environmental Medicine, SE-171 77, Karolinska Institutet, Stockholm, Sweden; Department of Medical Epidemiology and Biostatistics, SE-171 77, Karolinska Institutet, Stockholm, Sweden; Department of Medical Epidemiology and Biostatistics, Karolinska Institutet, SE-171 77, Stockholm, Sweden; Center of Public Health Sciences, Faculty of Medicine, University of Iceland, IS-101 Reykjavík, Iceland; Department of Biostatistics and Epidemiology, School of Public Health and Health Sciences, University of Massachusetts Amherst, Amherst, MA, USA; Department of Health Promotion and Policy, School of Public Health and Health Sciences, University of Massachusetts Amherst, Amherst, MA, USA

## Abstract

**Importance:** Premenstrual disorders (PMDs) are prevalent and significantly impair women’s quality of life. Yet, the long-term impact on work capacity is not well understood.

**Objective:** To prospectively examine the association between PMDs and work incapacity, including sick leave and unemployment.

**Setting, Design and Participants:** A prospective cohort study of 15,857 women aged 15-60 years and employed at baseline in the LifeGene Study with linkage to population and health registers in Sweden.

**Exposure:** PMDs were identified via register-based clinical diagnoses and symptom questionnaires.

**Main Outcomes and Measures:** We extracted information on sick leave and unemployment status from national registers. We used Poisson regression to estimate incidence rate ratios (IRRs) of sick leave and unemployment comparing women with PMDs to those without.

**Results:** In total, 2,585 (16.30%) women (mean age 32.52 years) reported symptom burden indicative of probable a PMD. With a median follow-up of 9.17 years, 6,741 (42.51%) and 1,485 (9.36%) women were exposed to at least one sick leave and unemployment during follow-up, respectively. Compared to women without PMDs, those with PMDs had 40% and 27% higher risks of sick leave (fully adjusted-IRR 1.40, 95% CI 1.31-1.49) and unemployment (IRR 1.27, 95% CI 1.10-1.46), respectively. The association was particularly stronger for long-term sick leave (≥ 90 days) (IRR 1.69, 95% CI 1.50-1.91), and sick leave due to depression (IRR 1.41, 95% CI 1.27-1.56). In addition, comparable associations for sick leave and unemployment were yielded, when comparing women with and without a history of depression/anxiety.

**Conclusions:** Employed women with PMDs are at increased risk of sick leave and unemployment, underscoring the potential long-term health and socioeconomic consequences of this prevalent condition. Improved clinical management of comorbidities and workplace policies are needed to support women affected by PMDs.

## Introduction

Premenstrual disorders (PMDs), including premenstrual syndrome (PMS) affects 20-30% of women of reproductive age of which premenstrual dysphoric disorder (PMDD) impacts 2.1-6.4%^1–3^ with debilitating psychological and physical symptoms typically emerging before menses.^3^ These symptoms recur with each menstrual cycle and significantly impair women’s quality of life throughout the reproductive years.^4^ Moreover, PMDs can have far-reaching effects on health. We have illustrated that women with PMDs face negative health consequences ranging from mental disorders, such as perinatal depression^5^, suicidal attempt^6^, and death of suicide^7^ to somatic conditions, such as, early menopause and severe perimenopause symptoms^8^, hypertension^9^, cardiovascular diseases^7^, and autoimmune diseases (personal communication). Meanwhile, moderate-to-severe PMS as well as PMDD significantly increase the burden of healthcare expenditures for patients^10^.

Although many women with PMDs received their diagnosis in their 30s, a previous study by our group suggests that about 70% of them experience symptom onset before age 20.^11^ For many, PMDs may considerably affect women’s working life from early adulthood. PMDs have recently been associated with women’s ability to work, including^12^ decreased work performance and reduced work productivity.^12–17^ It is therefore plausible that PMDs are associated with more severe work incapacity, such as increased rates of sick leave and job loss, either due to PMDs or related health consequences.

However, only a few studies have investigated the relationship between PMDs and sick leave. A cross-sectional study showed that up to 16% of women with PMDs had sick leave during the past year.^18^ Another study from the U.S. reported that women with PMS experienced reduced productivity and increased absenteeism (odds ratio 2.2; 95% CI 1.2–4.0) compared to women without, over a short follow-up of two menstrual cycles.^19^ Prospective data with adequate follow-up are however needed to fully understand the impact of PMDs and related health consequences on sick leave. Moreover, we are not aware of any study investigating the influence of PMDs on loss of job. Here, leveraging a prospective cohort of women employed at baseline in Sweden, we aimed to examine the risk of sick leave and unemployment (as a proxy for job loss) among women with PMDs, compared to women without PMDs.

## METHODS

### Data source

LifeGene is a longitudinal population-based cohort in Sweden.^20^ Briefly, in 2009, residents in Sweden aged between 18 and 50 years were recruited through simple random sampling from internet survey, with subsequent snowball sampling and self-registration online. Participants provided data on history of diseases and related risk factors (e.g., lifestyle) between 2009 and 2019.^21^ More than 52,000 participants underwent survey data collection^22^ and a total of 24,332 women responded to the baseline questionnaire, with 78% of them residing in Stockholm. Form of consent was obtained upon first access of the survey.^20^

We linked women to multiple Swedish registers through their unique personal identification number.^23,24^ The Longitudinal Integration Database for Health Insurance and Labor Market (LISA) collects information on all employee’s education, income and occupation since 1990.^25^ The National Social Insurance Agency’s Micro Data for Analyses of the Social Insurance (MiDAS) records data on sick leave (longer than 14 days for all employees^26^ and shorter leaves for self-employees^27^). The National Patient Register (NPR) contains data on all hospitalization diagnoses since 1987 and specialist care-based outpatient diagnoses since 2001.^28^ The Swedish Prescribed Drug Register (SPDR) records filled prescriptions from all pharmacies in Sweden since July 2005.^29^ Primary care register from Stockholm County records clinical diagnoses made by general practitioner.

The study was approved by the Swedish Ethical Review Authority, Sweden (2021-02775).

### Study design

We conducted a prospective cohort study of women of reproductive age nested within the LifeGene. Among 24,332 female participants, we excluded women aged <15 or >60 years, and those without employment status at baseline according to LISA (**Figure 1**). Finally, we included 15,857 women for analysis. This study followed the Strengthening the Reporting of Observational Studies in Epidemiology (STROBE) reporting guideline.

**Figure 1.**
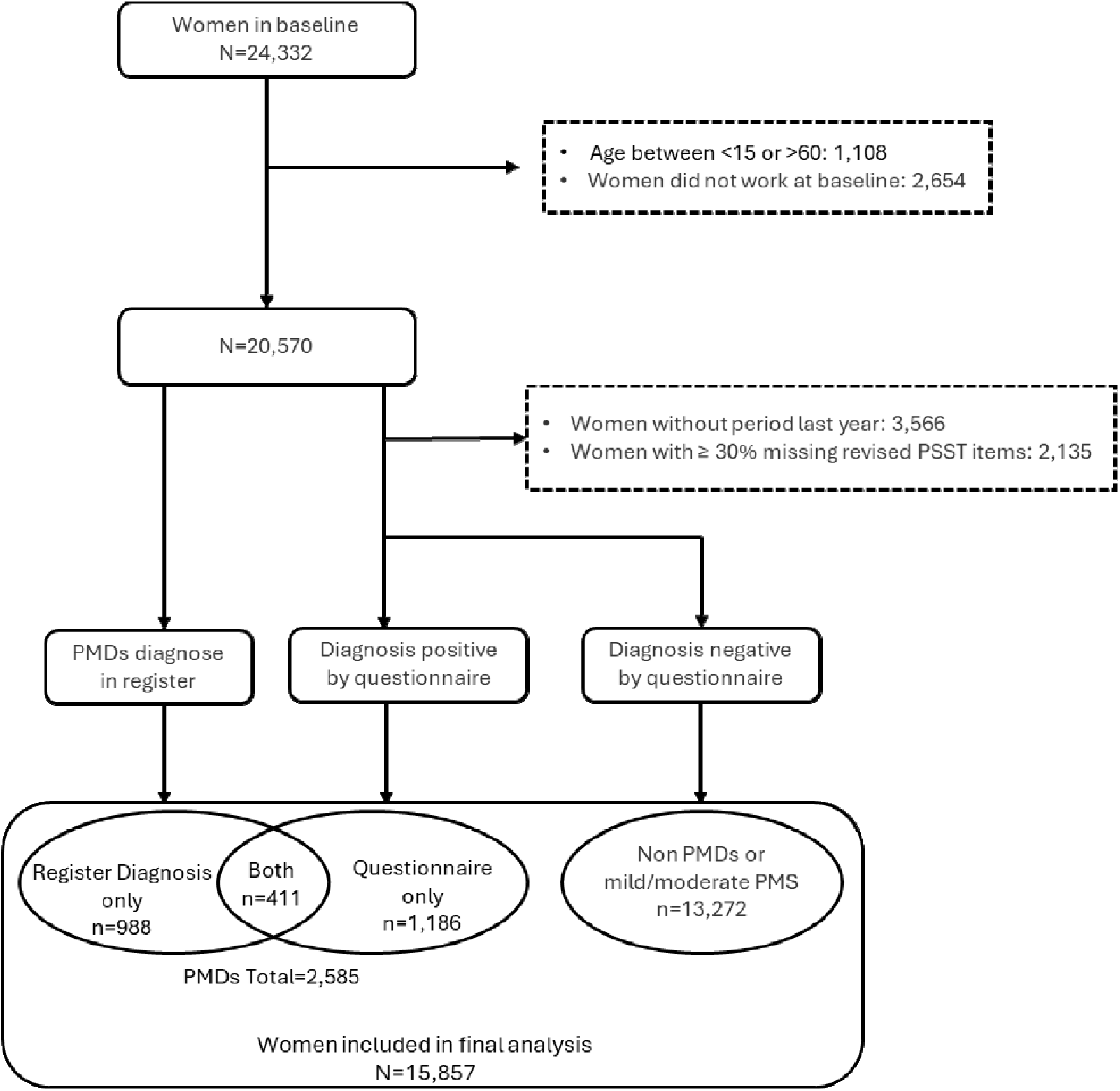
Flowchart. Abbreviation: PSST, Premenstrual Symptoms Screening Tool; N, number; PMDs, premenstrual disorder.

### Assessment of PMDs

We identified PMDs using clinical diagnoses indicated in healthcare registers both before baseline and during follow-up, and *via* questionnaire-based assessment administered at baseline. For the latter assessment, we excluded women without menstrual cycles in the year prior to baseline, and those who missed at least 30% of items in PMDs assessment questionnaire. A participant was considered to have a PMDs if indicated in either source. As described elsewhere,^7^ in register data, we identified clinical diagnoses of PMDs from the NPR (specialist care) and primary care register from Stockholm County, using International Classification of Diseases, Tenth Revision (ICD-10) code (N943, 625E). To complement the diagnoses made in primary care in counties other than Stockholm, we also identified PMDs using prescriptions of serotonin reuptake inhibitors (SSRIs; ATC code: N06AB) and other antidepressants (ATC code: N06AA, N06AB, N06AX) or hormonal contraceptives (ATC code: G03A, G02B) indicating a PMDs diagnosis or a specific dosing for PMDs (intermittent use of antidepressants) (Table S1).

A modified version of Premenstrual Symptoms Screening Tool (PSST)^30^ was used to assess PMDs at baseline. As described elsewhere,^31^ upon confirming three screening questions about symptom timing and impact on psychosocial functioning, participants rated the severity of 15 physical, behavioral, and emotional symptoms based on Diagnostic and Statistical Manual of Mental Disorders, Fifth Edition (DSM-V) criteria^32^ from 0 (none) to 3 (severe). Probable PMDs cases were categorized as follows: (1) one or more moderate/severe affective symptoms; (2) four or more other moderate/severe symptoms; and (3) at least one symptom with moderate/severe impact on relationships or social activities. Probable PMDs cases were further classified as PMS or probable PMDD; the latter was identified per DSM-V^32^ criteria: (1) one or more severe affective symptoms; and (2) at least one symptom severely affecting relationships or social activities. The original PSST has been validated in other populations with the sensitivity ranged from 77.4% to 90.9%.^33–35^

### Ascertainment of sick leave and unemployment

Information on sick leave was extracted from the MiDAS. Under the Swedish Sick Pay Act, employers are required to report to the National Social Insurance Agency when an employee has more than 15 days of sick leave. Length of sick leave was recorded in MiDAS. Such information was used to classify short-term (less than 90 days) and long-term (90 days or more) sick leave. In addition, we obtained diagnoses underlying sick leave, based on certificates from the physicians for each sick leave episode.

Employment status was extracted from a LISA annual report variable, gainful employment status, based on a) the presence of an income statement in November of the year in question, and b) a minimum yearly income (different levels by year, age and sex). Self-employed individuals are considered employed regardless of their registered salary, as long as their company is recorded as active.^25^ The status was recorded as “employment”, “temporary employment” and “unemployment”. The former two were deemed as employment in this study.

### Covariates

Information on covariates was derived at baseline. Data on age, country of birth, civil status, parity, smoking, alcohol consumption, adverse childhood experiences (ACEs) including parental divorce, abuse by family member, non-sexual assault by someone known, and sexual victimization, were collected through questionnaire, while body mass index (BMI) was derived from self-reported weight and height. Since depression and anxiety are common comorbidities of PMDs,^1,36,37^ we obtained diagnoses of depression and anxiety from registers (Table S1). In addition, educational level^38^ at baseline was retrieved from LISA.

### Statistical analysis

Women were followed from the baseline until the first occurrence of sick leave/ unemployment, death as identified from The National Cause of Death Register (CDR),^39^ emigration as obtained from the Total Population Register, age 67 (recommended age at retirement in Sweden),^40^ or December 31, 2020, whichever occurred first. The follow-up of unexposed women was additionally censored if they received a diagnosis of PMDs and thereafter they contributed to the follow-up of the exposed group.

Poisson regression was used to estimate incidence rate ratios (IRR) with 95% confidence intervals (CIs) of sick leave and unemployment by comparing women with and without PMDs, respectively. IRR was also estimated for the PMDs subtypes (PMS and PMDD) based on questionnaire assessment. Two models were developed to address potential confounding, including demographics,^41,42^ socioeconomic status,^42,43^ and lifestyle.^44,45^ In model 1, age was adjusted for. In model 2, BMI, civil status, ACEs, education level, and country of birth, parity, smoking, and alcohol consumption were additionally adjusted for. Model 2 was selected as the primary model for analysis in this study.

To shed light on the severity and nature of sick leave, we estimated the association for short-term and long-term sick leave separately, and for the ten most common diagnosis categories underlying sick leave.

We performed several additional analyses. First, to explore potential risk modification, we performed stratified analysis by psychiatric comorbidities (depression/anxiety) and history of sick leave (in the analysis of sick leave). Second, to account for multiple events during follow-up, we estimated the associations for recurrent sick leaves and unemployment status. Finally, although prospective symptom charting is recommended for PMDs diagnosis in the Swedish clinical guidelines,^46^ we lacked such information in registers. Therefore, we defined PMDs as those confirmed by both register-recorded diagnoses and questionnaire-based assessments, likely improving the validity of PMDs ascertainment.

The register data were prepared in SAS statistical software version 9.4 (SAS Institute, Cary, NC), while other data preparation and statistical analyses were performed in R software, version 4.0.5. Data were analyzed from January 18, 2023, to May 5, 2024.

## Results

At a mean age of 32.52 (SD 8.45) years, 2,585 (16.30%) women had either a register-recorded or questionnaire-assessed PMDs from baseline (n=1,879) or reported during follow-up (n=706). Compared to women without PMDs, those with PMDs were older and more likely to endorse ACEs, have been born abroad, have given birth, and have smoked in the past. Women with PMDs had a slightly higher mean BMI (all P<0.05; Table 1). Moreover, they were more likely to report a diagnosis of depression or anxiety.

**Table 1.** Characteristics of women with premenstrual disorder (PMDs) and without PMDs, N (%).

|  | No PMDs<br>N=13,272 | PMDs<br>N=2,585 | P |
| --- | --- | --- | --- |
| **Age (mean (SD))** | 32.37 (8.5) | 33.31 (8.1) | <0.001 |
| 15-19 | 367 (2.8) | 33 (1.3) | <0.001 |
| 20-29 | 5,511 (41.5) | 948 (36.7) |  |
| 30-39 | 4,339 (32.7) | 976 (37.8) |  |
| 40-49 | 2,652 (20.0) | 550 (21.3) |  |
| 50-60 | 403 (3.0) | 78 (3.0) |  |
| **BMI, kg/m2** |  |  | <0.001 |
| <18.5 | 420 (3.2) | 54 (2.1) |  |
| 18.5-25 | 9,939 (74.9) | 1,902 (73.6) |  |
| 25.1-30 | 1,701 (12.8) | 319 (12.3) |  |
| >30 | 319 (2.4) | 59 (2.3) |  |
| Unknown | 893 (6.7) | 251 (9.7) |  |
| **Civil status** |  |  | <0.001 |
| Non-cohabitation | 5,187 (39.1) | 907 (35.1) |  |
| Cohabitation | 7,852 (59.2) | 1,588 (61.4) |  |
| Unknown | 233 (1.8) | 90 (3.5) |  |
| **ACEs** |  |  | <0.001 |
| No | 8,549 (64.4) | 1,423 (55.0) |  |
| Yes | 4,710 (35.5) | 1,088 (42.1) |  |
| Unknown | 13 (0.1) | 74 (2.9) |  |
| **Education Level** |  |  | 0.014 |
| Primary school | 603 (4.5) | 88 (3.4) |  |
| Secondary school | 3,390 (25.5) | 691 (26.7) |  |
| Post-secondary | 9,223 (69.5) | 1,801 (69.7) |  |
| Other/Unknown | 56 (0.4) | 5 (0.2) |  |
| **Country of birth** |  |  | <0.001 |
| Sweden | 12,020 (90.6) | 2,279 (88.2) |  |
| Not Sweden | 1,252 (9.4) | 306 (11.8) |  |
| **Parity** |  |  | <0.001 |
| 0 | 8,601 (64.8) | 1,510 (58.4) |  |
| 1-2 | 3,748 (28.2) | 806 (31.2) |  |
| 3 | 771 (5.8) | 188 (7.3) |  |
| Unknown | 152 (1.1) | 81 (3.1) |  |
| **Smoking status** |  |  | <0.001 |
| Never | 4,673 (35.2) | 656 (25.4) |  |
| Past | 7,522 (56.7) | 1,611 (62.3) |  |
| Current | 1,031 (7.8) | 244 (9.4) |  |
| Unknown | 46 (0.3) | 74 (2.9) |  |
| **Alcohol consumption** |  |  | <0.001 |
| Never | 410 (3.1) | 67 (2.6) |  |
| 1-3 times a month or less often | 6,963 (52.5) | 1,331 (51.5) |  |
| More than once a week | 5,685 (42.8) | 1,085 (42.0) |  |
| Unknown | 214 (1.6) | 102 (3.9) |  |
| **Depression<sup>a</sup>** |  |  | <0.001 |
| No | 11,202 (84.4) | 1,728 (66.8) |  |
| Yes | 2,070 (15.6) | 857 (33.2) |  |
| **Anxiety<sup>a</sup>** |  |  | <0.001 |
| No | 11,392 (85.8) | 1,956 (75.7) |  |
| Yes | 1,880 (14.2) | 629 (24.3) |  |
| **History of Sick leave** |  |  | <0.001 |
| No | 3,503 (56.8) | 763 (50.5) |  |
| Yes | 2,659 (43.2) | 747 (49.5) |  |

Abbreviation: BMI, body mass index (kg/m<sup>2</sup>); ACEs, adverse childhood experiences; N, number; %, percentage.
<sup>a</sup> depression and anxiety diagnosis from registers

### Risks of sick leave and unemployment

During a median follow-up of 9.17 years (mean age at end of follow-up 40.70 (SD 8.87) years), 6,741 (42.51%) and 1,485 (9.36%) women had a sick leave and unemployment recorded in registers, respectively. Compared to unaffected women, women with PMDs had 40% (IRR 1.40, 95% CI 1.31-1.49) and 27% (IRR 1.27, 95%CI 1.10-1.46) higher risks of sick leave and unemployment, respectively (Table 2, Model 2). Given that the questionnaire assessment was primarily designed to screen PMDD, 1,489 (57.60%) of all PMDs cases were classified as PMDD and 108 (4.18%) were identified as severe PMS. Statistically comparable associations for sick leave and unemployment were observed between PMDs and PMDD. While no increased association of sick leave and unemployment were found in women with severe PMS.

**Table 2.** Incidence Rate Ratios (IRRs) and 95% confidence intervals (CIs) of unemployment and sick leave among women with premenstrual disorders (PMDs) compared to those without.

|  | N of events (IR<sup>a</sup>) | Model 1<sup>b</sup><br>IRR (95% CI) | Model 2<sup>c</sup><br>IRR (95% CI) |
| --- | --- | --- | --- |
| **Sick Leave** |  |  |  |
| No PMDs | 5,623 (65.60) | 1.00 | 1.00 |
| PMDs | 1,118 (96.85) | 1.45 (1.36-1.55) | 1.40 (1.31-1.49) |
| -PMS<sup>d</sup> | 47 (70.95) | 1.07 (0.80-1.42) | 1.05 (0.79-1.40) |
| -PMDD<sup>d</sup> | 801 (94.97) | 1.43 (1.33-1.54) | 1.38 (1.28-1.49) |
| **Unemployment** |  |  |  |
| No PMDs | 1,251 (11.58) | 1.00 | 1.00 |
| PMDs | 234 (14.06) | 1.33 (1.15-1.53) | 1.27 (1.10-1.46) |
| -PMS<sup>d</sup> | 10 (11.39) | 1.00 (0.54-1.87) | 0.97 (0.52-1.81) |
| -PMDD<sup>d</sup> | 172 (14.45) | 1.35 (1.15-1.58) | 1.29 (1.10-1.52) |

Abbreviations: N, number; IR, incidence rate; IRR, Incidence rate ratios; CI, confidence interval; PMDs, premenstrual disorders; PMS, premenstrual syndrome; PMDD, premenstrual dysphoric disorder.
<sup>a</sup> Per 1000 person-years, unadjusted.
<sup>b</sup> Estimates were adjusted for age.
<sup>c</sup> Estimates were additionally adjusted for BMI, civil status, ACEs, education level, and country of birth, parity, smoking, and alcohol assumption.
<sup>d</sup> In the analysis of PMDs subtype, only questionnaire-assessed PMDs were included because ICD code cannot be used to distinguish PMDD from PMS. Specifically, 1,597 women with PMDs (108 PMS and 1,489 PMDD) were included in the analyses for sick leave and unemployment.

### Subtypes of sick leave

When analyzing length of sick leave, a stronger association was suggested for long-term sick leave (IRR 1.69, 95% CI 1.50-1.91) than short-term (IRR 1.35, 95% CI 1.25-1.46) sick leave (Table 3, Model 2). In the analysis of underlying causes of sick leave, the strongest association was noted for sick leave due to depression (IRR 1.41, 95% CI 1.27-1.56), post-traumatic stress disorder (PTSD) (IRR 1.40, 95% CI 1.28-1.54), followed by pregnancy complications (IRR 1.36, 95% CI 1.24-1.49) (Table 4).

**Table 3.** Incidence Rate Ratios (IRRs) and 95% confidence intervals (CIs) for Short-Term and Long-Term Sick Leave among women with premenstrual disorder (PMDs) compared to women without PMDs.

|  | N of events (IR <sup>a</sup>) | Model 1<sup>b</sup> | Model 2<sup>c</sup> |
| --- | --- | --- | --- |
|  |  | IRR (95% CI) | IRR (95% CI) |
| **Short term<sup>d</sup>** |  |  |  |
| No PMDs | 4,142 (52.00) | 1.00 | 1.00 |
| PMDs | 767 (73.77) | 1.40 (1.29-1.51) | 1.35 (1.25-1.46) |
| **Long term<sup>e</sup>** |  |  |  |
| No PMDs | 1,481 (21.43) | 1.00 | 1.00 |
| PMDs | 351 (39.43) | 1.79 (1.59-2.01) | 1.69 (1.50-1.91) |

Abbreviations: N, number; IR, incidence rate; IRR, Incidence rate ratios; CI, confidence interval; PMDs, premenstrual disorder.
<sup>a</sup> Per 1000 person-years, unadjusted.
<sup>b</sup> Estimates were adjusted for age.
<sup>c</sup> Estimates were additionally adjusted for BMI, civil status, ACEs, education level, and country of birth, parity, smoking, and alcohol assumption.
<sup>d</sup> Short term: The days of sick leave was less than 90 days.
<sup>e</sup> Long term: The days of sick leave was greater than or equal to 90 days.

**Table 4.**
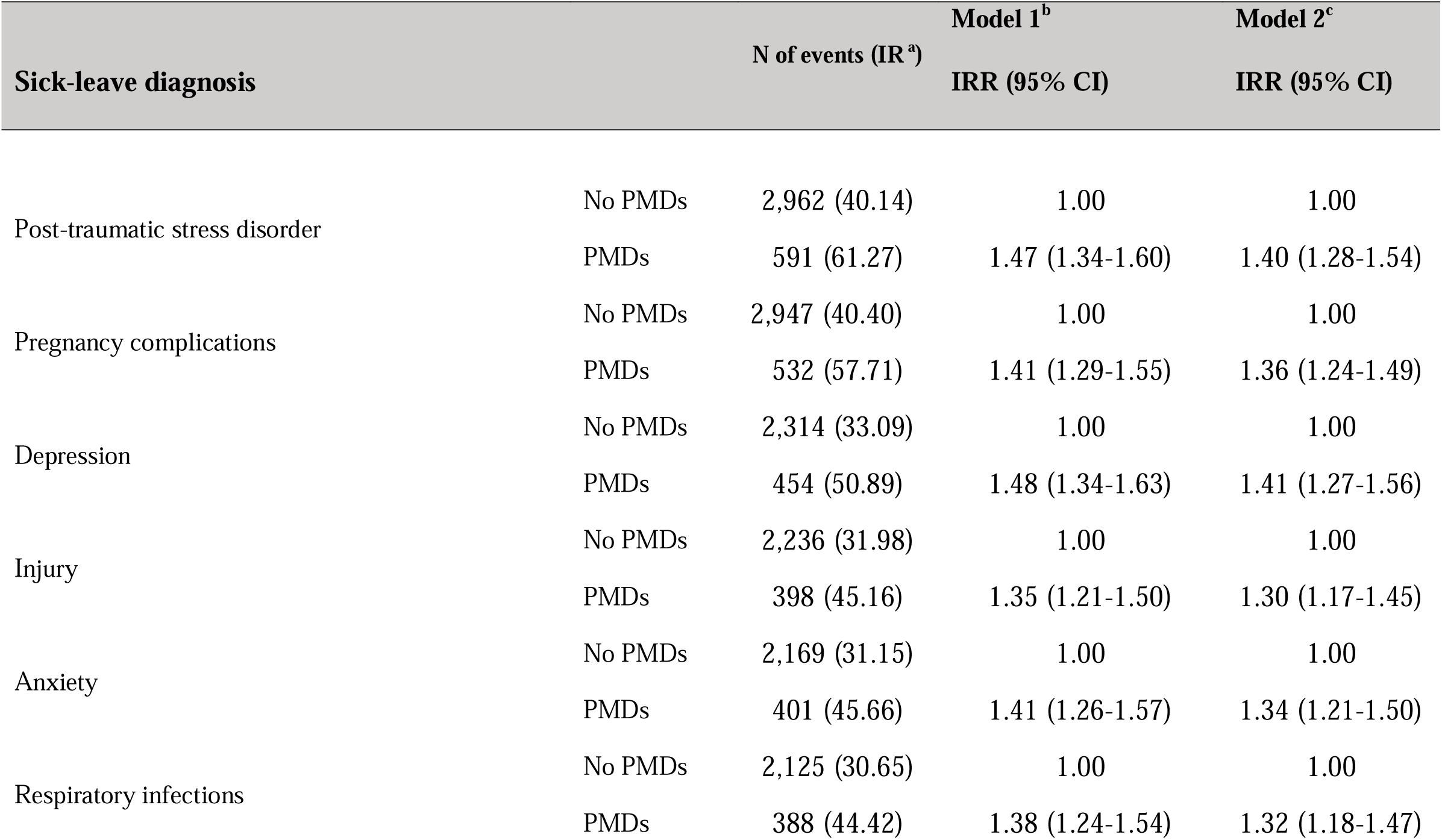
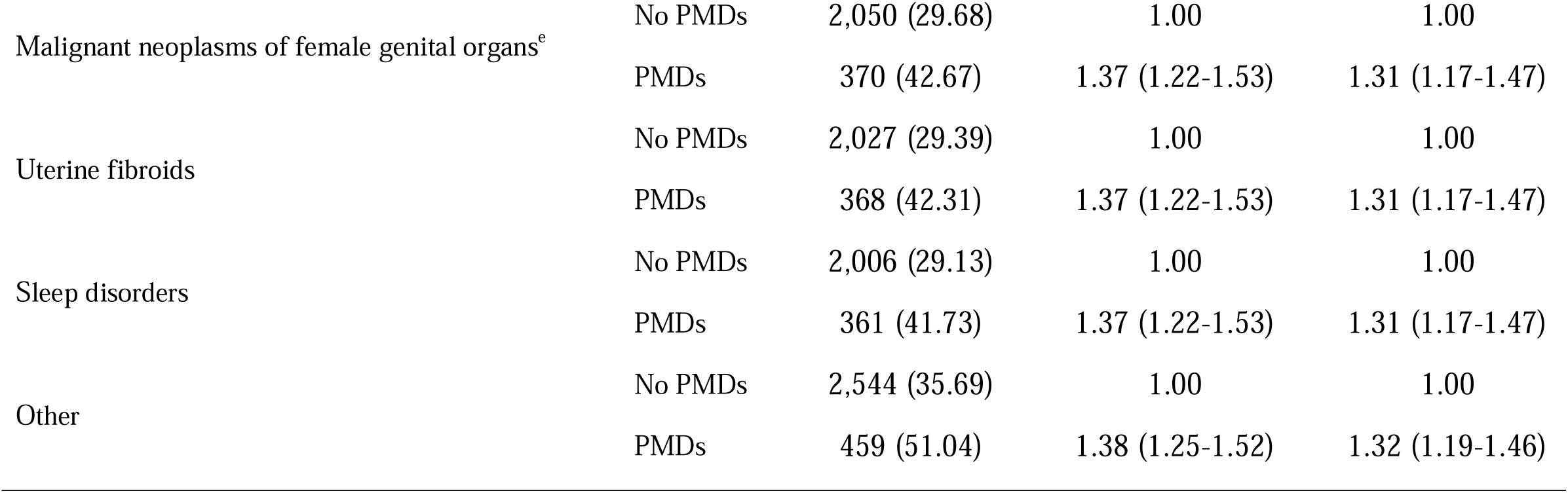
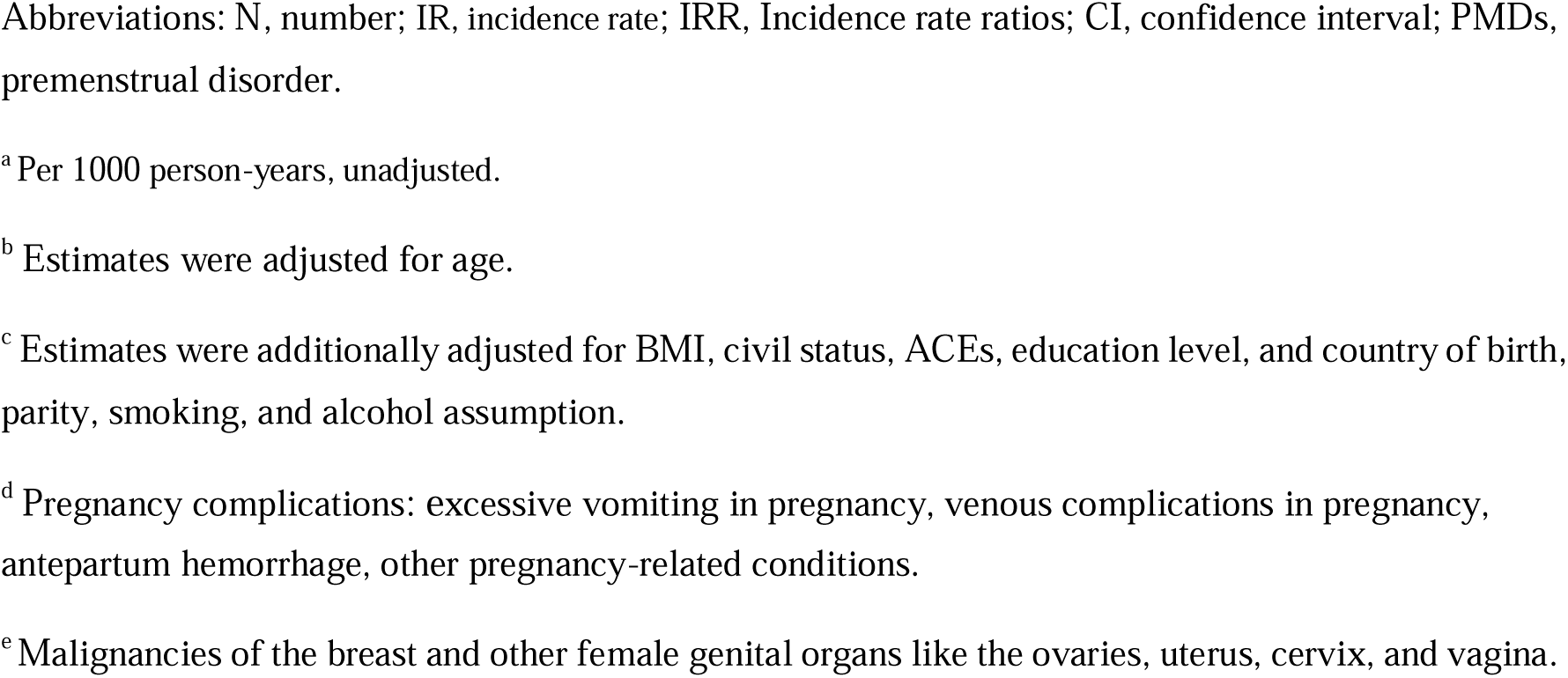
Incidence rate ratios (IRRs) with 95% confidence intervals (CIs) for the diagnoses underlying sick leave among women with premenstrual disorder (PMDs) compared to women without PMDs.

| Sick-leave diagnosis |  | N of events (IR <sup>a</sup>) | Model 1<sup>b</sup> | Model 2<sup>c</sup> |
| --- | --- | --- | --- | --- |
|  |  |  | IRR (95% CI) | IRR (95% CI) |
| Post-traumatic stress disorder | No PMDs | 2,962 (40.14) | 1.00 | 1.00 |
|  | PMDs | 591 (61.27) | 1.47 (1.34-1.60) | 1.40 (1.28-1.54) |
| Pregnancy complications | No PMDs | 2,947 (40.40) | 1.00 | 1.00 |
|  | PMDs | 532 (57.71) | 1.41 (1.29-1.55) | 1.36 (1.24-1.49) |
| Depression | No PMDs | 2,314 (33.09) | 1.00 | 1.00 |
|  | PMDs | 454 (50.89) | 1.48 (1.34-1.63) | 1.41 (1.27-1.56) |
| Injury | No PMDs | 2,236 (31.98) | 1.00 | 1.00 |
|  | PMDs | 398 (45.16) | 1.35 (1.21-1.50) | 1.30 (1.17-1.45) |
| Anxiety | No PMDs | 2,169 (31.15) | 1.00 | 1.00 |
|  | PMDs | 401 (45.66) | 1.41 (1.26-1.57) | 1.34 (1.21-1.50) |
| Respiratory infections | No PMDs | 2,125 (30.65) | 1.00 | 1.00 |
|  | PMDs | 388 (44.42) | 1.38 (1.24-1.54) | 1.32 (1.18-1.47) |
| Malignant neoplasms of female genital organs<sup>e</sup> | No PMDs | 2,050 (29.68) | 1.00 | 1.00 |
|  | PMDs | 370 (42.67) | 1.37 (1.22-1.53) | 1.31 (1.17-1.47) |
| Uterine fibroids | No PMDs | 2,027 (29.39) | 1.00 | 1.00 |
|  | PMDs | 368 (42.31) | 1.37 (1.22-1.53) | 1.31 (1.17-1.47) |
| Sleep disorders | No PMDs | 2,006 (29.13) | 1.00 | 1.00 |
|  | PMDs | 361 (41.73) | 1.37 (1.22-1.53) | 1.31 (1.17-1.47) |
| Other | No PMDs | 2,544 (35.69) | 1.00 | 1.00 |
|  | PMDs | 459 (51.04) | 1.38 (1.25-1.52) | 1.32 (1.19-1.46) |

Abbreviations: N, number; IR, incidence rate; IRR, Incidence rate ratios; CI, confidence interval; PMDs, premenstrual disorder.
<sup>a</sup> Per 1000 person-years, unadjusted.
<sup>b</sup> Estimates were adjusted for age.
<sup>c</sup> Estimates were additionally adjusted for BMI, civil status, ACEs, education level, and country of birth, parity, smoking, and alcohol assumption.
<sup>d</sup> Pregnancy complications: excessive vomiting in pregnancy, venous complications in pregnancy, antepartum hemorrhage, other pregnancy-related conditions.
<sup>e</sup> Malignancies of the breast and other female genital organs like the ovaries, uterus, cervix, and vagina.

### Additional analysis

We observed comparable associations for sick leave and unemployment, when comparing women with and without history of depression and anxiety (Table S2, S3). Further, similar association with sick leave was found between women with and without history of sick leave (Table S4). In addition, largely comparable results were yielded when analyzing multiple events of unemployment/sick leave during follow-up (Table S5).

We identified 411 (15.90%) women with PMDs confirmed by both register diagnosis and questionnaire assessment (Figure 1). Compared to the main analysis, the association was seemingly stronger for sick leave (IRR 1.97, 95% CI 1.63-2.40) whereas the association for unemployment (IRR 1.28, 95% CI 0.95-1.73) was similar but not statistically significant (Table S6).

## Discussion

In this prospective cohort study of 15,857 employed women with a maximum follow-up of 12 years, we found that women with PMDs, mainly PMDD, are at a higher risk of both sick leave and unemployment. Women with PMDs also have a greater risk increase for long-term sick leave and are more likely to take sick leave due to depression, PTSD, and pregnancy complications. This study marks the first comprehensive assessment of the association of PMDs on sick leave and unemployment, providing important insights into the long-term health and socioeconomic impact entailing PMDs.

Previous studies reported that PMDs affect work productivity and increase work absenteeism.^12,13^ However, many previous studies focused on specific occupations, which may have a limited generalizability to other occupations.^15,16,47^ Additionally, prior reports primarily comprised cross-sectional studies^15,47–50^ or prospective studies with short follow-up. ^19,51–53^ The only research on sick leave^18^ was focused on short sick leave (less than 14 days), ignoring the chronic nature of PMDs and the contribution to sick leave might be last beyond 14 days. Our study found that over a long follow-up period, women with PMDs had a 40% higher risk of being on sick leave for more than two weeks, compared to unaffected women. However, we found no elevated association of sick leave among women with PMS. The increased risk of sick leave might be related to their premenstrual symptoms.^54,55^ Women with PMDD might struggle more with managing emotional and physical symptoms, such as severe mood swings, anxiety or tension, and breast tenderness or swelling, which can interfere with their daily tasks, leading to frequent absences and decreased performance.^56^ Such impact is typically confined within the week of late luteal phase.^57^ Indeed, in our data, no sick leave (≥14 days) was directly prescribed for underlying diagnosis of PMDs (ICD-10 N94.3). However, the recurrent nature of PMDs^58^ may have a cumulative effect on work performance and the relationship with co-workers, which can contribute to adverse working-related outcomes in the long run.^53,59,60^ While more studies are warranted to illustrate the reasons underlying an association between PMDs and sick leave. Indeed, our study has illustrated the higher risk when accounting for repeated sick leave, and a stronger association with long-term sick leave (90 days or more).

Moreover, our findings may be explained by comorbidities associated with PMDs. Indeed, we found women with PMDs were more likely to take sick leave due to depression and PTSD compared to unaffected women. It is well-documented that women with PMDs are likely to have psychiatric comorbidities, such as depression, anxiety, and PTSD.^61–64^ In addition, it has been shown that women with PMDs experience severe physical and psychological symptoms in early pregnancy,^65^ while our previous study found a higher risk of antepartum and postpartum depression among women with PMDs.^5^ In this study, we noted a positive association between PMDs and sick leave due to pregnancy complications. Future research is needed to understand PMD-associated pregnancy conditions and corresponding sick leaves. It is worth noting that we found moderate difference in elevated association across underlying reasons for sick leave. This may be due to competing risk, i.e., a patient may have multiple conditions/comorbidities, but only the condition leading to the first sick leave was accounted for.

A cross-sectional study among Japanese nurses showed that symptoms during menstruation can increase willingness to quit a job.^66^ To our knowledge, no study has investigated the association of PMDs with unemployment. Here, we found that women with PMDD had 30% higher risk of unemployment relative to unaffected women. Such risk can be attributed to the cumulative effect of recurrent PMDD episodes as well as the chronic comorbidities. It is important to note that in our study, we were unable to determine whether the unemployment was due to voluntary termination or to being fired, which requires further investigation.

Our study is a prospective cohort study with adequate length of and complete follow-up, including participants from the general population, and comprehensive adjustment for potential confounders. However, several limitations should be considered. First, the probable PMDs identified from questionnaire assessment were based on retrospective symptom reporting, which could introduce potential misclassification. However, since participants were not aware of their future working outcomes, such misclassification was likely non-differential and would have attenuated the results towards null. Indeed, a stronger association with sick leave was observed when restricting PMDs cases confirmed by both clinical diagnosis and questionnaire assessment, assuming a better validity of diagnosis. Moreover, given the nature of our data sources, mild/moderate PMS was not captured in the present study. Our findings may not apply for the group with milder symptoms. Second, MiDAS did not include sick leave less than 15 days except for self-employees. Thus, our results may not be generalized to sick leave with shorter days. Third, compared to the Swedish general population, LifeGene participants are younger and have higher socio-economic status^21^. However, we have accounted for these potential selection forces in our analyses, and we found comparable PMDs^67^ and sick leave^68^ prevalence among the general population and Lifegene participants. In addition, we cannot exclude the possibility of reverse causation, namely PMDs can be the consequence of some chronic conditions and related sick leave. For instance, isolation and reduced activity levels due to sick leave can exacerbate mental health problems,^69^ and unemployment can lead to increased symptoms of somatization, depression and anxiety.^70^ However, we have observed comparable results regardless of depression history. Finally, our results may not generalize to countries with different labor market and welfare policies.

In conclusion, our study suggests that employed women with PMDs are at a higher risk of sick leave and unemployment. These findings highlight the substantial health and socioeconomic burden associated with PMDs, emphasizing the need for effective clinical management of PMDs and comorbidities as well as society and workplace support systems for affected women.

## Data sharing

Due to privacy protection measures, such as the General Data Protection Regulation (GDPR), the cohort’s information is not publicly accessible. Access to Lifegene resources is granted only after an ethical evaluation by the relevant authorities (https://ki.se/en/research/research-infrastructure-and-environments/core-facilities-for-researc h/ki-biobank-core-facility-kibb/lifegene). For further information on acquiring access to Swedish register data, consult the Swedish National Board of Health and Welfare’s website (https://bestalladata.socialstyrelsen.se/) and/or the Statistics Sweden website (https://www.scb.se/vara-tjanster/bestall-data-och-statistik/)

## Data Availability

Due to privacy protection measures, such as the General Data Protection Regulation (GDPR), the cohort's information is not publicly accessible. Access to Lifegene resources is granted only after an ethical evaluation by the relevant authorities (https://ki.se/en/research/research-infrastructure-and-environments/core-facilities-for-research/ki-biobank-core-facility-kibb/lifegene). For further information on acquiring access to Swedish register data, consult the Swedish National Board of Health and Welfare's website (https://bestalladata.socialstyrelsen.se/) and/or the Statistics Sweden website (https://www.scb.se/vara-tjanster/bestall-data-och-statistik/)

https://bestalladata.socialstyrelsen.se/

https://www.scb.se/vara-tjanster/bestall-data-och-statistik/

