## Supplementary material for "Premenstrual disorders and risk of sick leave and unemployment: a prospective cohort study of 15,857 women in Sweden": Table S1 Table S2 Table S3 Table S4 Table S5 Table S6

**Table S1. Codes for identification of premenstrual disorders**

**Table S2. Incidence rate ratios (IRRs) with 95% confidence intervals (CIs) of sick leave among women with premenstrual disorders (PMDs) compared to women without PMDs stratified by diagnosis of depression and anxiety separately.**

**Table S3. Incidence rate ratios (IRRs) with 95% confidence intervals (CIs) of unemployment among women with premenstrual disorders (PMDs) compared to women without PMDs stratified by diagnosis of depression and anxiety separately.**

**Table S4. Incidence rate ratios (IRRs) with 95% confidence intervals (CIs) of sick leave among women with premenstrual disorders (PMDs) compared to women without PMDs stratified by history of sick leave.**

**Table S5. Incidence rate ratios (IRRs) with 95% confidence intervals (CIs) of recurring work-related outcomes among women with premenstrual disorders (PMDs) compared to women without PMDs.**

**Table S6. Incidence rate ratios (IRRs) with 95% confidence intervals (CIs) of sick leave and unemployment among women with premenstrual disorders (PMDs) confirmed by both clinical diagnosis and questionnaire assessment, compared to women without PMDs.**

**Table S1. Codes for identification of premenstrual disorders**

| **Diagnosis** | **Source** | **Type** | **Codes** |
| --- | --- | --- | --- |
| Premenstrual disorders | NPR | ICD-10 | N943, 625E |
|  | SPDR | ATC | N06AA, N06AB, N06AX, G03A, G02B |
|  |  |  | With a written indication for PMDs in Swedish:  "PMS", "PREMENSTRUELLT SYNDROM", "PREMENSTRUELLT DYSFORSIKT SYNDROM", "PREMENSTRUELLT DYSFORI", "PMD", "PMDD", "PMDS", "MENS" |
| Depression | NPR | ICD-10 | F32-F33 |
|  | SPDR | ATC | N06A |
| Anxiety | NPR | ICD-10 | F40-F41 |
|  | SPDR | ATC | N05B |

Abbreviations: NPR, The National Patient Register; SPDR, The Swedish Prescribed Drug Register; ICD, International Classification of Diseases; ATC, Anatomical Therapeutic Chemical; PMDs, Premenstrual disorders.

**Table S2. Incidence rate ratios (IRRs) with 95% confidence intervals (CIs) of sick leave among women with premenstrual disorders (PMDs) compared to women without PMDs stratified by diagnosis of depression and anxiety separately.**

|  | **N (IR ^a^)** | **Model 1^b^** | **Model 2^c^** |
| --- | --- | --- | --- |
|  |  | **IRR (95% CI)** | **IRR (95% CI)** |
| *Depression diagnosis* | |  |  |
| **without** |  |  |  |
| No PMDs | 4,400 (58.37) | 1.00 | 1.00 |
| PMDs | 635 (77.61) | 1.32 (1.21-1.44) | 1.29 (1.18-1.40) |
| **with** |  |  |  |
| No PMDs | 1,223 (118.36) | 1.00 | 1.00 |
| PMDs | 483 (143.67) | 1.20 (1.08-1.34) | 1.19 (1.07-1.33) |
| *Anxiety diagnosis* | |  |  |
| **without** |  |  |  |
| No PMDs | 4,602 (60.33) | 1.00 | 1.00 |
| PMDs | 781 (85.02) | 1.39 (1.29-1.50) | 1.35 (1.25-1.46) |
| **with** |  |  |  |
| No PMDs | 1,021 (108.26) | 1.00 | 1.00 |
| PMDs | 337 (142.96) | 1.30 (1.15-1.47) | 1.29 (1.13-1.46) |

Abbreviations: N, number of events; IR, incidence rate; IRR, Incidence rate ratios; CI, confidence interval; PMDs, premenstrual disorder.

^a^ Per 1000 person-years, unadjusted.

^b^ Estimates were adjusted for age.

^c^ Estimates were additionally adjusted for BMI, civil status, ACEs, education level, country of birth, parity, smoke, alcohol assumption.

**Table S3. Incidence rate ratios (IRRs) with 95% confidence intervals (CIs) of unemployment among women with premenstrual disorders (PMDs) compared to women without PMDs stratified by diagnosis of depression and anxiety separately.**

|  | **N (IR ^a^)** | **Model 1^b^** | **Model 2^c^** |
| --- | --- | --- | --- |
|  |  | **IRR (95% CI)** | **IRR (95% CI)** |
| *Depression diagnosis* | |  |  |
| **without** |  |  |  |
| No PMDs | 981 (10.61) | 1.00 | 1.00 |
| PMDs | 143 (12.98) | 1.3 (1.09-1.54) | 1.23 (1.03-1.47) |
| **with** |  |  |  |
| No PMDs | 270 (17.35) | 1.00 | 1.00 |
| PMDs | 91 (16.19) | 1.02 (0.80-1.3) | 1.06 (0.82-1.36) |
| *Anxiety diagnosis* | |  |  |
| **without** |  |  |  |
| No PMDs | 1008 (10.68) | 1.00 | 1.00 |
| PMDs | 164 (12.81) | 1.32 (1.12-1.56) | 1.27 (1.08-1.51) |
| **with** |  |  |  |
| No PMDs | 243 (17.76) | 1.00 | 1.00 |
| PMDs | 70 (18.23) | 1.09 (0.83-1.42) | 1.06 (0.80-1.40) |

Abbreviations: N, number of events; IR, incidence rate; IRR, Incidence rate ratios; CI, confidence interval; PMDs, premenstrual disorder.

^a^ Per 1000 person-years, unadjusted.

^b^ Estimates were adjusted for age.

^c^ Estimates were additionally adjusted for BMI, civil status, ACEs, education level, country of birth, parity, smoke, alcohol assumption.

**Table S4. Incidence rate ratios (IRRs) with 95% confidence intervals (CIs) of sick leave among women with premenstrual disorders (PMDs) compared to women without PMDs stratified by history of sick leave.**

|  | **N (IR ^a^)** | | **Model 1^b^** | **Model 2^c^** |
| --- | --- | --- | --- | --- |
|  |  | | **IRR (95% CI)** | **IRR (95% CI)** |
| *History of sick leave* | |  | |  |
| **without** |  | |  |  |
| No PMDs | 3,133 (189.17) | | 1.00 | 1.00 |
| PMDs | 535 (228.77) | | 1.19 (1.09-1.30) | 1.17 (1.07-1.29) |
| **with** |  | |  |  |
| No PMDs | 2,490 (253.12) | | 1.00 | 1.00 |
| PMDs | 583 (281.47) | | 1.11 (1.02-1.22) | 1.09 (0.99-1.19) |

Abbreviations: N, number of events; IR, incidence rate; IRR, Incidence rate ratios; CI, confidence interval; PMDs, premenstrual disorder.

^a^ Per 1000 person-years, unadjusted.

^b^ Estimates were adjusted for age.

^c^ Estimates were additionally adjusted for BMI, civil status, ACEs, education level, country of birth, parity, smoke, alcohol assumption.

**Table S5. Incidence rate ratios (IRRs) with 95% confidence intervals (CIs) of recurring work-related outcomes among women with premenstrual disorders (PMDs) compared to women without PMDs.**

|  | **N (IR ^a^)** | | **Model 1^b^** | **Model 2^c^** |
| --- | --- | --- | --- | --- |
|  |  | | **IRR (95% CI)** | **IRR (95% CI)** |
| **Unemployment** | |  | | |
| No PMDs | 2,511 (17.84) | | 1.00 | 1.00 |
| PMDs | 507 (21.94) | | 1.32 (1.20-1.45) | 1.27 (1.15-1.4) |
| **Sick leave** | |  | | |
| **Total counts** |  | |  |  |
| No PMDs | 13,066 (92.83) | | 1.00 | 1.00 |
| PMDs | 2,838 (122.82) | | 1.3 (1.25-1.36) | 1.24 (1.19-1.29) |
| **Short-term counts^d^** |  | |  |  |
| No PMDs | 9,768 (69.4) | | 1.00 | 1.00 |
| PMDs | 1,998 (86.47) | | 1.23 (1.17-1.29) | 1.17 (1.11-1.23) |
| **Long-term counts^e^** |  | |  |  |
| No PMDs | 3,298 (23.43) | | 1.00 | 1.00 |
| PMDs | 840 (36.35) | | 1.52 (1.41-1.64) | 1.45 (1.34-1.57) |

Abbreviations: N, number of recurring work-related events; IR, incidence rate; IRR, Incidence rate ratios; CI, confidence interval; PMDs, premenstrual disorder.

^a^ Per 1000 person-years, unadjusted.

^b^ Estimates were adjusted for age.

^c^ Estimates were additionally adjusted for BMI, civil status, ACEs, education level, country of birth, parity, smoke, alcohol assumption.

^d^ Total number of episodes for sick leave was less than 90 days.

^e^ Total number of episodes for sick leave was greater than or equal to 90 days.

**Table S6. Incidence rate ratios (IRRs) with 95% confidence intervals (CIs) of sick leave and unemployment among women with premenstrual disorders (PMDs) confirmed by both clinical diagnosis and questionnaire assessment, compared to women without PMDs.**

|  | **N (IR ^a^)** | | **Model 1^b^** | **Model 2^c^** |
| --- | --- | --- | --- | --- |
|  |  | | **IRR (95% CI)** | **IRR (95% CI)** |
| **Sick leave** | |  | | |
| No PMDs | 5,623 (65.6) | | 1.00 | 1.00 |
| PMDs | 105 (137.96) | | 1.99 (1.64-2.41) | 1.97 (1.63-2.40) |
| **Unemployment** | |  | | |
| No PMDs | 1,251 (11.58) | | 1.00 | 1.00 |
| PMDs | 44 (13.36) | | 1.30 (0.96-1.76) | 1.28 (0.95-1.73) |

Abbreviations: N, number of events; IR, incidence rate; IRR, Incidence rate ratios; CI, confidence interval; PMDs, premenstrual disorder.

^a^ Per 1000 person-years, unadjusted.

^b^ Estimates were adjusted for age.

^c^ Estimates were additionally adjusted for BMI, civil status, ACEs, education level, country of birth, parity, smoke, alcohol assumption.
